# Surveillance of COVID-19 vaccination in US nursing homes, December 2020-April 2021

**DOI:** 10.1101/2021.05.14.21257224

**Authors:** Andrew Isaac Geller, Daniel S. Budnitz, Heather Dubendris, Radhika Gharpure, Minn Minn Soe, Hsiu Wu, Elizabeth J. Kalayil, Andrea L. Benin, Suchita A. Patel, Megan C. Lindley, Ruth Link-Gelles

## Abstract

Monitoring COVID-19 vaccination coverage among nursing home (NH) residents and staff is important to ensure high coverage and guide patient-safety policies. With the termination of the federal Pharmacy Partnership for Long-Term Care Program, another source of facility-based vaccination data is needed. We compared numbers of COVID-19 vaccinations administered to NH residents and staff reported by pharmacies participating in the temporary federal Pharmacy Partnership for Long-Term Care Program with those reported by NHs participating in new COVID-19 vaccination modules of CDC’s National Healthcare Safety Network (NHSN). Pearson correlation coefficients comparing the number vaccinated between the two approaches were 0.89, 0.96, and 0.97 for residents and 0.74, 0.90, and 0.90 for staff, in the weeks ending January 3, 10, and 17, respectively. Based on subsequent NHSN reporting, vaccination coverage with ≥1 vaccine dose reached 77% for residents and 50% for staff the week ending January 31 and plateaued through April 2021.

**Three-question summary box:**

1. *What is the current understanding of the subject?* Because of high risk of disease, nursing home residents and staff were prioritized for COVID-19 vaccination when doses were limited.
2. *What does this report add to the literature?* National monitoring of nursing home residents and staff vaccination coverage through the CDC National Healthcare Safety Network (NHSN) correlated with vaccination administration reports from the federal Pharmacy Partnership for Long-Term Care Program in January 2021. NHSN-reported vaccination coverage rates plateaued from February through April 2021.
3. What are the implications for public health practice? NHSN can track COVID-19 vaccination in nursing homes and help guide efforts to increase vaccine uptake in residents and staff.

## Introduction

Residents and staff of nursing homes (NHs) are at increased risk for SARS-CoV-2 infection,^1-3^ and NH residents are at risk for severe complications of COVID-19.^1,4-7^ To help protect this disproportionately affected population, in December 2020, the Centers for Disease Control and Prevention (CDC) launched the Pharmacy Partnership for Long-Term Care Program, facilitating on-site COVID-19 vaccination of NH residents as well as NH staff.^8^ Among the approximately 11,000 NHs that had at least one on-site vaccination clinic in the first month of the program, an estimated three-quarters of residents (77.8%) and one-third of staff (37.5%) were vaccinated with ≥1 dose of a COVID-19 vaccine.^9^

CDC’s National Healthcare Safety Network (NHSN) is the nation’s web-based surveillance program for monitoring healthcare-associated adverse events, annual healthcare staff influenza vaccinations, and other patient-safety practices. In December 2020, NHSN released COVID-19 reporting modules for tracking vaccination coverage among residents and staff of long-term care facilities.^10^ These modules allow for voluntary weekly reporting by facilities and are designed to collect data on the number of current residents and staff eligible for vaccination and who have been vaccinated.^10^

The Pharmacy Partnership for Long-Term Care Program was designed as a time-limited initiative to facilitate a limited number of vaccination clinics at each participating facility. With the conclusion of the program, reporting of facility-level resident and staff COVID-19 vaccine administration data to CDC has ceased, but the need for a national data source for ongoing monitoring of COVID-19 vaccination in NHs remains, so the Centers for Medicare and Medicaid Services has begun to require NH reporting through NHSN as of June 2021.^11^

The objectives of this analysis were to: 1) compare numbers of COVID-19 vaccinations administered to NH residents and staff in January 2021 as reported by pharmacies participating in the Pharmacy Partnership for Long-Term Care Program, and the NHSN COVID-19 vaccination reporting modules, among CMS-certified NHs that participated in both; and 2) report weekly COVID-19 vaccination coverage for residents and staff among CMS-certified NHs voluntarily reporting through NHSN from December 2020 through April 2021.

## Methods

### Correlation of COVID-19 Vaccination Administration Reporting

We used facility NHSN Organization ID and CMS Certification Numbers to identify NHs that had their first Pharmacy Partnership for Long-Term Care Program vaccination clinic in the weeks ending January 3, 10, and 17, and also reported vaccination coverage data through NHSN. Data were accessed on February 22, 2021. We identified a minority number of facilities in which the total number of doses administered to residents or staff according to Pharmacy Partnership for Long-Term Care Program reporting exceeded the total number of current residents or staff eligible to be vaccinated as reported to NHSN, and set the total vaccinated to the total number of residents or staff reported to NHSN. We calculated Pearson correlation coefficients to compare vaccine administration data reported by the Pharmacy Partnership for Long-Term Care Program and NHSN.

### COVID-19 Vaccination Coverage in Facilities Reporting to NHSN

Among all CMS-certified NHs reporting vaccination data through NHSN, we calculated the numbers and proportions of residents and staff who received doses of COVID-19 vaccine among facilities reporting each week from the week ending December 20, 2020 through the week ending May 9, 2021.

This activity was reviewed by CDC and was conducted consistent with applicable federal law and CDC policy.^§^

## Results

### Correlation of COVID-19 Vaccination Administration Reporting

During the three consecutive one-week periods, 1028 CMS-certified NHs reported vaccination data on residents to both the Pharmacy Partnership for Long-Term Care Program as well as to NHSN; 818 CMS-certified NHs participated in the Pharmacy Partnership for Long-Term Care Program and reported COVID-19 vaccination coverage to NHSN for staff. Among these NHs, the number of vaccine doses administered in the weeks ending January 3, 10, and 17, 2021 exceeded the total number of current residents eligible to receive COVID-19 vaccination as reported to NHSN for 62 (17.7%), 53 (12.5%), and 33 (12.9%) facilities, respectively, and the number of vaccine doses administered exceeded the total number of eligible staff reported to NHSN for 13 (4.5%), 9 (2.7%), and 5 (2.6%) facilities, respectively.

Pearson correlation coefficients for numbers of COVID-19 vaccinations administered to NH residents and staff as reported by the Pharmacy Partnership for Long-Term Care Program and the NHSN COVID-19 vaccination coverage modules were 0.89, 0.96, and 0.97 for NH residents and 0.74, 0.90, and 0.90 for NH staff, in the weeks ending January 3, 10, and 17, 2021.

### COVID-19 Vaccination Coverage in Facilities Reporting to NHSN

A total of 5065 NHs reported resident COVID-19 vaccination coverage for at least 1 week from the week ending December 20, 2020 through the week ending May 9, 2021; a total of 3554 NHs reported staff COVID-19 vaccination coverage for at least 1 week during the same period. Among NHs reporting to NHSN, coverage with at least one dose of COVID-19 vaccine rose rapidly and reached 77.0% for residents (**Figure 1**) and 49.6% for staff (**Figure 2**) during the week ending January 31, 2021. Coverage with at least one dose of COVID-19 vaccine then plateaued for both NH residents and NH staff through April 2021. By the week ending January 31, 2021, 50.5% of residents and 30.5% of staff had received a complete series of COVID-19 vaccine, and the proportion of residents and staff who had received a complete series continued to increase through April 2021.

**Figure 1.**
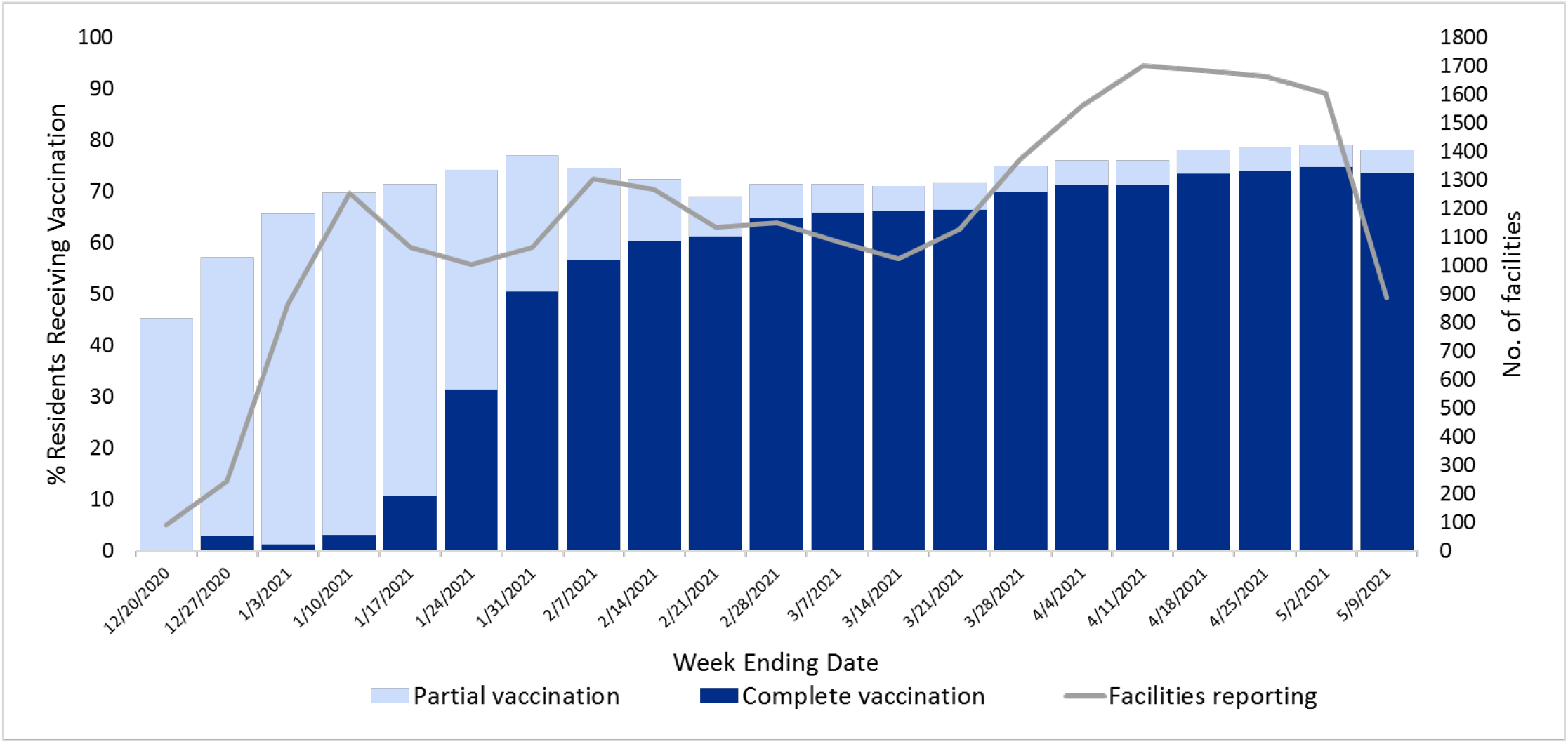
COVID-19 vaccination coverage of nursing home residents among facilities reporting to the National Healthcare Safety Network each week. Shaded bars indicate the proportion of residents receiving partial and complete vaccination, and line plot indicates the number of facilities reporting resident vaccination coverage. Data are current as of May 11, 2021. Abbreviation: COVID-19, coronavirus disease 2019.

**Figure 2.**
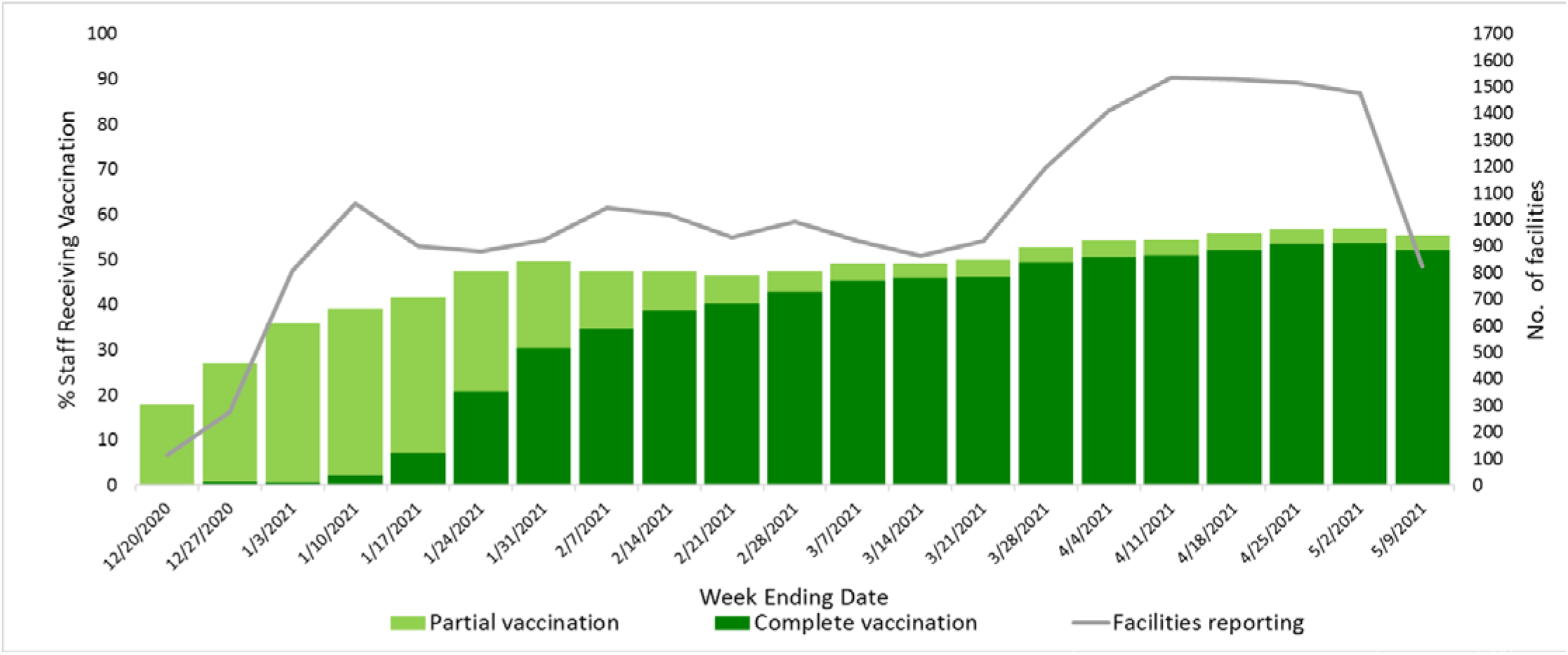
COVID-19 vaccination coverage of nursing home staff among facilities reporting to the National Healthcare Safety Network each week. Shaded bars indicate the proportion of staff receiving partial and complete vaccination, and line plot indicates the number of facilities reporting staff vaccination coverage. Data are current as of May 11, 2021. Abbreviation: COVID-19, coronavirus disease 2019.

In the most recent 4-week period analyzed, from April 12, 2021–May 9, 2021, 2,276 NHs reported resident vaccination data, representing 13.8% of all CMS-certified NHs enrolled in NHSN, and 1,997 NHs reported staff vaccination data, representing 12.1% of NHs. These NHs reported 77.1% of residents (n=130,427) and 55.5% of staff (n=131,317) received at least partial vaccination; 72.8% (n=123,131) and 51.8% (n=122,545) received a completed vaccination series.

## Discussion

High correlation between vaccine administration reported by the Pharmacy Partnership for Long-Term Care Program and NHSN COVID-19 vaccination reporting modules suggests NHSN can be used to track COVID-19 vaccination in NHs. With the completion of the Pharmacy Partnership for Long-Term Care Program, there remains a public health need to track COVID-19 vaccination coverage in NHs, where residents remain at elevated risk for morbidity and mortality and both residents as well as staff are at high risk for SARS-CoV-2 acquisition and transmission.

Continued tracking of COVID-19 vaccination coverage in NHs is important for several reasons. First, vaccination coverage of staff members has plateaued, suggesting further efforts are needed to increase coverage in these important healthcare personnel. Historically, high staff turnover rates at NHs— approximating 100 percent annually^12^—warrant ongoing COVID-19 vaccination efforts. Additionally, addressing vaccine hesitancy of NH staff^13-15^ and improving vaccine acceptance is needed. Ongoing weekly surveillance from NHSN can track the impact of such efforts. Second, although currently plateaued at a rate higher than in staff members, NH resident vaccination coverage should continue to be monitored to track national vaccination efforts, as new residents are admitted to facilities on a daily basis. Third, jurisdictions can use weekly coverage rates to target intervention resources to facilities with greatest need of vaccination uptake.^16^ Fourth, NHs may use facility vaccination rates to guide infection control policies, such as those pertaining to indoor visitation with NH residents.^17^

This report is subject to a few limitations. First, correlation between Pharmacy Partnership for Long-Term Care Program and NHSN data is not expected to be exact. Pharmacy Partnership for Long-Term Care Program data include only individuals vaccinated on-site, whereas NHSN counts persons vaccinated at the facility or elsewhere. Second, there were instances in which the total number of doses administered to residents or staff according to Pharmacy Partnership for Long-Term Care Program reporting exceeded the total number of current residents or staff reported to NHSN. One explanation is that the number of vaccine doses brought to a facility exceeded the number of residents and staff who decided to be vaccinated, and to avoid wastage other persons (e.g., healthcare workers from other facilities) were vaccinated and recorded as facility residents or staff. Another possible explanation is that the number of residents or staff were underreported to NHSN. Nonetheless, the number of facilities reporting excess doses administered was small and declined each week. Third, facilities reporting NHSN COVID-19 vaccination coverage may not be representative of all nursing homes. Reporting to NHSN was voluntary, and approximately one-tenth of eligible facilities reported vaccination coverage data in the last 4 weeks of the study period. Nonetheless, the January vaccination coverage rates reported by NHSN the Pharmacy Partnership for Long-Term Care Program were similar for residents (77.3% versus 77.8%) and staff (50.0% vs. 37.5%)^9^.

As the Pharmacy Partnership for Long-Term Care Program has been replaced with direct federal allocations of COVID-19 vaccine to the long-term care pharmacies, which have traditionally provided vaccines to nursing homes,^8^ NHSN is now the single national source of facility-level resident and staff COVID-19 vaccination data to monitor vaccination progress, target vaccination support programs, and help inform infection control policies.

## Supporting information

Data for Figures

## Data Availability

Data supporting the findings of the study are found in the manuscript and/or supplementary files. Any other data can be furnished upon request.

## Acknowledgments

The authors would like to thank the following people for assistance with data analysis and data quality assurance: Karen Jones and Audrey Robnett-Brown of the Centers for Disease Control and Prevention (CDC); Dorothy Dennard, Shiyi Wang, Tianyi Zhou, and Adewole Adetokun (of Goldbelt C6, LLC, contractor to CDC); and Emily Witt (of CACI, Inc, contractor to CDC).

## Disclaimer

The findings and conclusions in this report are those of the authors and do not necessarily represent the official position of the Centers for Disease Control and Prevention.

## Footnotes

§ See e.g., 45 C.F.R. part 46, 21 C.F.R. part 56; 42 U.S.C. §241(d); 5 U.S.C. §552a; 44 U.S.C. §3501 et seq.

